# Community transmission of Mpox clade Ib not driven through sexual exposures in Uvira, eastern Democratic Republic of Congo, June – October 2024

**DOI:** 10.1101/2025.02.25.25322371

**Authors:** Patrick Musole Bugeme, Patrick Kazuba Bugale, Trust Faraja Mukika, Megan O’Driscoll, Javier Perez-Saez, Levi Bugwaja, Salomon Mashupe Shangula, Willy Kasi, Justin Bengehya, Stephanie Ngai, Antonio Isidro Carrion Martin, Jules Jackson, Patrick Katoto, Esto Bahizire, Noella Mulopo-Mukanya, Justin Lessler, Jackie Knee, Pauline Vetter, Elizabeth C. Lee, Daniel Mukadi-Bamuleka, Andrew S Azman, Espoir Bwenge Malembaka

**Affiliations:** Department of Epidemiology, Johns Hopkins University, Baltimore, MD 21205, USA; Center for Tropical Diseases and Global Health (CTDGH), Université Catholique de Bukavu, Bukavu, Democratic Republic of the Congo; Hôpital Général de Référence d’Uvira, Uvira Health Zone, Uvira, Democratic Republic of the Congo; Geneva Centre for Emerging Viral Diseases, Geneva University Hospitals, Geneva, Switzerland; Division Provinciale de la Santé Publique du Sud-Kivu, Bukavu, Democratic Republic of the Congo; Médecins Sans Frontières, Uvira, The Democratic Republic of the Congo; Médecins Sans Frontières, London, United Kingdom; Centre de Recherche en Sciences Naturelles de Lwiro, Bukavu, DR Congo; Rodolphe Merieux INRB-Goma Laboratory, Goma, Democratic Republic of Congo; University of North Carolina Population Center, University of North Carolina at Chapel Hill, Chapel Hill, NC, USA; Department of Epidemiology, Gillings School of Global Public Health, University of North Carolina at Chapel Hill, Chapel Hill, NC, USA; Department of Disease Control, London School of Hygiene & Tropical Medicine, London, UK; Department of Virology, Institut National de Recherche Biomédicale (INRB), Kinshasa, Democratic Republic of the Congo; Service de Microbiologie, Département de Biologie Médicale, Cliniques Universitaires de Kinshasa, Université de Kinshasa, Kinshasa, Democratic Republic of the Congo; Division of Tropical and Humanitarian Medicine, Geneva University Hospitals (HUG), Geneva, Switzerland

**Keywords:** mpox, clade 1b, children, household transmission

## Abstract

**Introduction:** In September 2023, mpox clade 1b emerged in Kamituga, a mining zone in South Kivu, DR Congo, primarily through sexual transmission. This study investigates an mpox clade 1b outbreak in Uvira, 300 km from Kamituga, where the first case was reported in May 2024.

**Methods:** From June-October 2024, clinical and household data from suspected mpox cases were collected at Uvira hospital and through investigations. Laboratory confirmation was performed via PCR. We analyzed demographic, exposure, and clinical data to characterize transmission patterns, severity, and risk factors.

**Results:** Among 972 suspected cases, 42.2% (411) had PCR results, with 78.1% positive. The median age was 9 years (IQR: 3-20), with 63.8% under 15 (n=621) and 32.1% under 5 (n=312). Severe cases (≥100 lesions) were more frequent in children under 15 (25.6%, 142/555) than older individuals (16.2%, 49/303; p<0.001). Acute malnutrition affected 8.6% (14/162) of children under 5. The case fatality ratio was 0.7% (7/972), rising to 4% (5/127) in infants under 1 year. Among HIV-tested individuals, 1.8% (6/329) were HIV-positive. Pregnant women accounted for 14.5% (19/131) of cases. Household exposures dominated (67.9%), with limited sexual (6%) or healthcare worker (0.4%) exposures. Female sex workers represented 1.2% of cases. Animal exposures were predominantly domestic (97.4%, 38/39).

**Conclusion:** This child-centered outbreak, driven by household transmission, underscores the need for pediatric vaccines, nutritional support, and household interventions. Adult-focused responses alone may be insufficient.

## Introduction

Monkeypox virus (MPXV) has been documented in humans in the Democratic Republic of the Congo (DRC) since 1970, historically causing small-scale zoonotic mpox outbreaks primarily affecting young children in the western and central provinces. These outbreaks were characterized by limited person-to-person transmission and with no significant differences in infection rates by sex.[1–3]

There are two distinct MPXV clades with marked geographic distribution, each comprising two subclades (a and b), and differing case fatality ratios influenced by factors such as clade virulence, host vulnerability, comorbidities, and access to care. Clade Ia, predominant in Central Africa and presumed more virulent, has a case fatality ratio (CFR) reaching 10%. [4,5] Clade IIa is more common west of the Dahomey Gap, from Ghana to Sierra Leone, with a CFR below 0.1%, while clade IIb predominates east of the Dahomey Gap in Nigeria and shows more variable CFRs, though generally lower than Clade I.[6,7] In September 2023, the South Kivu province of DRC reported its first-ever mpox outbreak in the remote mining region of Kamituga, caused by a newly-identified virus variant, clade Ib.[8] The virus rapidly spread across the Kivu region, Burundi, Rwanda, Uganda, Kenya, and beyond. This, combined with increased transmission of other mpox variants, mainly within the DRC but also in other African settings, prompted the World Health Organization (WHO) to declare it a Public Health Emergency of International Concern in August 2024.[9–11] Reports from Kamituga, where the clade Ib outbreak was first identified and where most patients were adults, suggested that sexual contact was the primary mode of transmission, and earlier studies in western DRC had already linked Clade I mpox infections to sexual exposure.[12] A cohort study conducted between May and October 2024 at Kamituga Mpox Treatment Center (MTC) found that 75% of all 510 suspected mpox cases (and 79% of 407 confirmed cases) were ≥15 years.[13] Notably, genital lesions were nearly three times more prevalent in patients ≥15 years (89%) than among children under 5 years of age (32%). Furthermore, 40% of patients with available contact information reported sexual exposure to a suspected or confirmed case. Another study conducted in the same area showed that nearly nine out of ten patients reported recent sexual contact in bars or hotels[14] However, official surveillance data suggested that as the outbreak spread to other health zones of South Kivu, the proportion of cases among children has increased, indicating a possible shift towards non-sexual transmission pathways in these areas.[15]

The first mpox cases in Uvira were officially reported on May 2, 2024, 8 months after the outbreak began in Kamituga. An MTC was opened at the Uvira General Hospital on June 8, 2024. Since June 17, 2024, a free mpox care package has been provided at the MTC with support from Médecins Sans Frontières, covering medical treatment for mpox and related complications, essential medications, meals for patients and caregivers.

Here, we provide new insights into the epidemiology, clinical features and risk factors associated with mpox clade Ib in Uvira, South Kivu, DRC, approximately 300 km from Kamituga.

## Methods

### Setting, design and population

We conducted a descriptive surveillance study of mpox in Uvira, a trading city on the northern shores of Lake Tanganyika, from June 3 to October 24, 2024. Uvira health zone is a mixture of rural and urban areas sharing land border with Burundi and connected to Tanzania and Zambia through the DRC’s second largest port. The city includes 18 of the 22 health areas of the Uvira health zone. There are an estimated 460,000 people in the Uvira health zone. Approximately 65% of the 460,000 people in the Uvira health zone live in the city (City of Uvira Official Population Count Data, 2024), with an estimated population density of 17,988 people per km^2^. The median household size in Uvira city is 8 individuals.[16,17] Agriculture, fisheries and trading play an important part in the local economy.[18]

Uvira is part of South Kivu Province, which, as of April 2024, hosted around 1.9 million of the 7 million people internally displaced in eastern DRC,[19] though exact official numbers per health zone are not readily available. The city also hosts refugees, mainly from Burundi, due to ongoing conflicts in the region. Uvira is also highly vulnerable to climate-related disasters. Between February and April 2024, flooding in Uvira and the neighboring Fizi territory submerged around 3,000 houses, disrupted three universities and five markets, and rendered 800 water taps nonfunctional, further exacerbating existing humanitarian needs.[19] [20]

### Clinical case definition and data collection

At the Uvira MTC, anyone presenting with a rash or skin lesions was considered a suspected mpox case and eligible for this analysis (Supplementary file). Suspected case basic data were recorded in MTC’s paper triage register which includes basic information, including patient identifiers, date of hospital admission or outpatient visit, symptom onset date, sex, age, profession, home address (health area and avenue), presence of fever, skin rash, or lesions on palms or soles, sample collection status, mpox laboratory result, hospitalization or outpatient management status, smallpox vaccination status, outcome details (dead, discharged, transferred) and date of outcome.

Two health zone physicians conducted clinical examinations and administered an in-depth electronic form via the KoboCollect platform, following WHO mpox surveillance guidelines.[21] This form captured all the paper register data along with additional details on clinical signs, symptoms and medical history, exposures and travel within the past 21 days, and care-seeking behavior. Vulnerability data, including pregnancy, breastfeeding, and child nutritional status, assessed using standard Mid-Upper Arm Circumference (MUAC) measurements,[22] were collected. A discharge form was also completed for each patient to record visit outcomes. Where residential addresses were available and located within the city, a team of three nurses visited the households of cases residing in the city to gather basic information on household composition and characteristics. Additionally, home visits were conducted in December 2024 for pregnant women residing within the city.

### Samples collection and testing

Skin lesions specimens were collected with a dry swab by trained MTC staff. When not possible, oropharyngeal sampling was collected in a viral transport medium. Samples were maintained at 4°C and transported to either the Rodolphe Mérieux INRB Goma Laboratory or the South Kivu Provincial Laboratory in Bukavu for testing (Supplementary File).

At the Rodolphe Mérieux INRB Goma Laboratory, samples were tested by qPCR with the Radi^®^ Fast Mpox Kits (KH Medical, Pyeongtaek-si, Gyeonggi-do, Republic of Korea) to detect MPXV Clades I and/or II. We considered a cycle threshold (Ct)-value of <40 threshold as positive. Additionally, a subset of samples was clade-typed with TIB Molbiol® qPCR kit (TIB Molbiol, Eresburgstrasse, Berlin, Germany).[23][24][23][24]

At the South Kivu Provincial Laboratory, samples were analyzed on the GeneXpert^®D^ platform (Xpert®□ Mpox, Cepheid, Sunnyvale, California, USA), designed to detect mpox DNA from MPXV Clade II and non-variola orthopoxvirus (OPX) genes in human specimens. Samples were classified as positive for MPXV Clade I when non-variola orthopoxvirus DNA was detected with no amplification of the Clade II MPXV target, following the manufacturer’s instructions.[25,26]

Whenever Rapid Diagnostic Tests (RDTs) were available, consenting hospitalized patients were screened for HIV and Syphilis using the SD Bioline^®^ HIV/Syphilis Duo Rapid Diagnostic Test (RDT) (Abbott Diagnostics, Gyeonggi-do, Republic of Korea). For patients testing positive for HIV on this RDT, two additional tests were performed with Stat-Pak (Chembio Diagnostic Systems, New York, USA) and Uni-Gold TM □ HIV (Trinity Biotech, Wicklow, Ireland) as per the HIV control program algorithm.

### Data management and analysis

We calculated the weekly number of clinically attended cases, by age group (< 1 year, 1-4 years, 5-14 years, 15-44 years and ≥45 years) and laboratory confirmation. We used WHO guidance to classify disease severity as mild (<25 skin lesions), moderate (25-99 lesions), severe (100-250 lesions), and very severe or grave (>250 lesions).[27] For children aged 3 to 59 months with MUAC measurements, we calculated MUAC-for-age (MFA) z-scores using the *zscorer* package in R.[28] Nutritional status was classified as normal (MFA ≥-2), moderate acute malnutrition (−3 ≤ MFA < -2), or severe acute malnutrition (MFA < -3).[22,29] We used data from four population-representative surveys conducted in Uvira city between 2021 and 2024 [17] (n=10604) to estimate the demographic structure of the population given that no census has been conducted in DRC since 1984. These data allowed us to compare the age distribution of suspected mpox cases to that of the general Uvira population, as well as to compare the age distribution of cases to that of their households, based on age composition collected during home visits. The characteristics of study participants were compared using the Wilcoxon rank sum, Pearson’s Chi-squared, or Fisher’s exact tests. All analyses were performed using R (4.4.1).

### Ethics considerations

For this study, we used the surveillance data from the Uvira health zone. Ethical approval to conduct this study was obtained from the Institutional Review Boards of the Johns Hopkins Bloomberg School of Public Health (reference number IRB00030442) and the Université Catholique de Bukavu (UCB/CIES/NC/019/2024).

## Results

### Socio-demographic and household characteristics

Between June 3 and October 24, 2024, 973 suspected mpox cases were recorded at the Uvira MTC, with 50.5% (n=491) being female. The median age of patients was 9 years (interquartile range [IQR]: 3–20 years). Most cases were children, with 63.7% (n=620) being under 15 years and 35.4% (n= 334) under 5 years (Table 1, Table S1, Figure 1). The proportion of cases under 15 years increased as the outbreak progressed until mid-July 2024 (Figure 1, Panel C). The age distribution of those with suspected mpox was younger than the overall Uvira population, with those 0–5 years old being particularly overrepresented (Figure 1, Panel A). Patients’ median household size was 8 (IQR: 5 – 10) people, with a median of 4 (IQR: 2.7 – 5.4) people per sleeping room (Table 1). Female sex workers made up 1.2% (n=12) of the total cases, and only four (0.4%) cases were healthcare workers (Table 1).

**Table 1.**
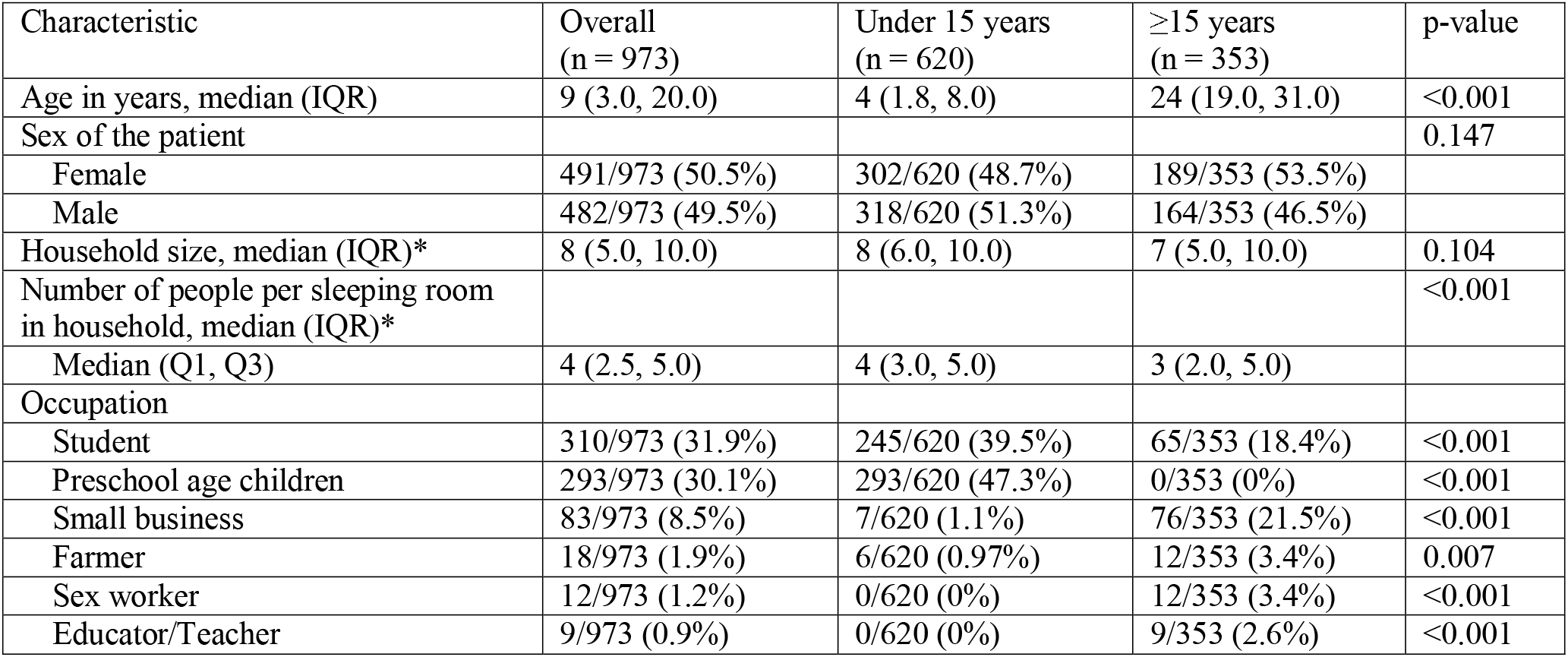
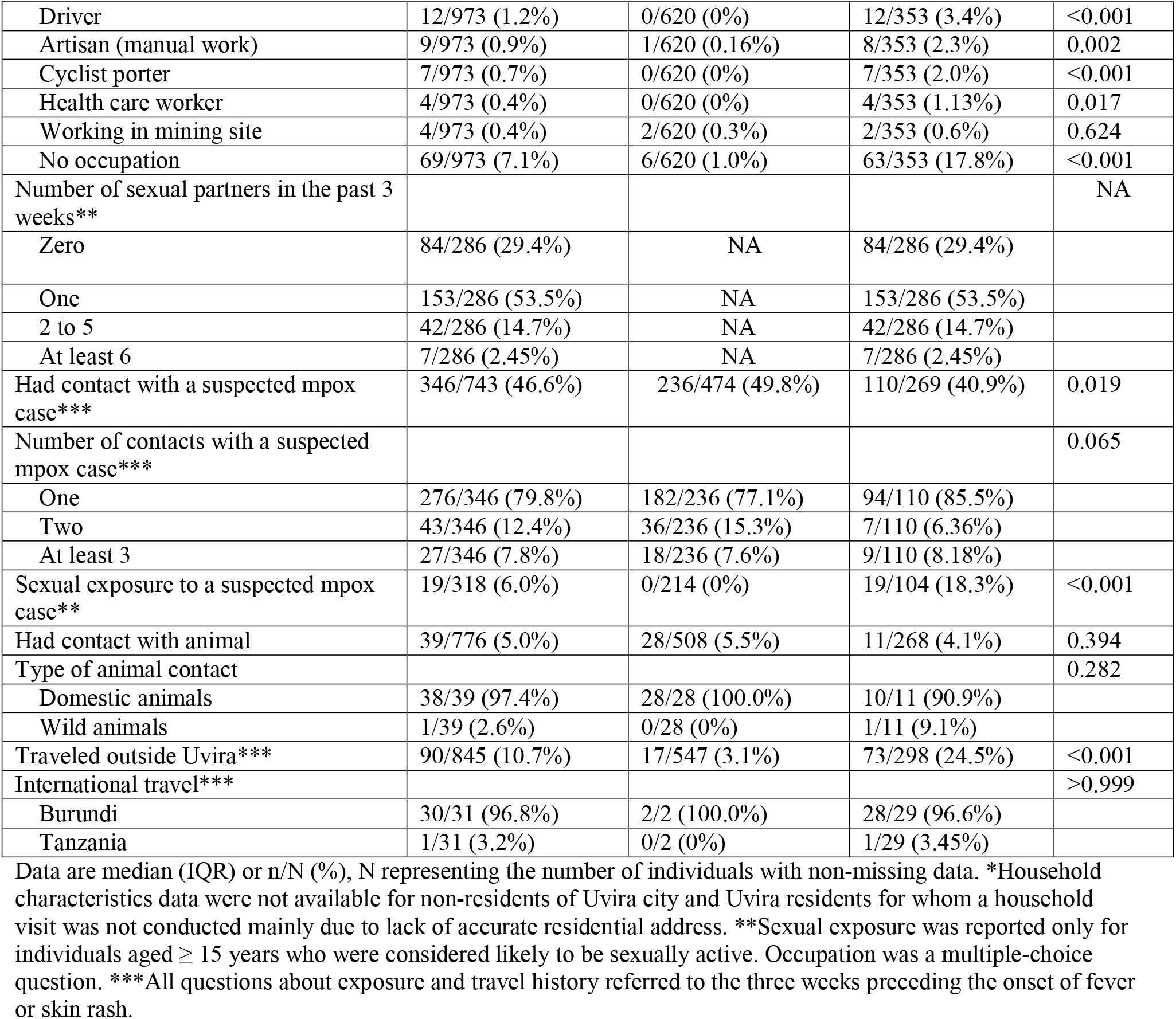
Socio-demographic characteristics and exposure history of suspected mpox cases seeking care at the Uvira Mpox Treatment Centre. Data are median (IQR) or n/N (%), N representing the number of individuals with non-missing data. ^*^ Household characteristics data were not available for non-residents of Uvira city and Uvira residents for whom a household visit was not conducted mainly due to lack of accurate residential address. ^**^ Sexual exposure was reported only for individuals aged ≥15 years who were considered likely to be sexually active. Occupation was a multiple-choice question. ^***^All questions about exposure and travel history referred to the three weeks preceding the onset of fever or skin rash.

**Figure 1.**
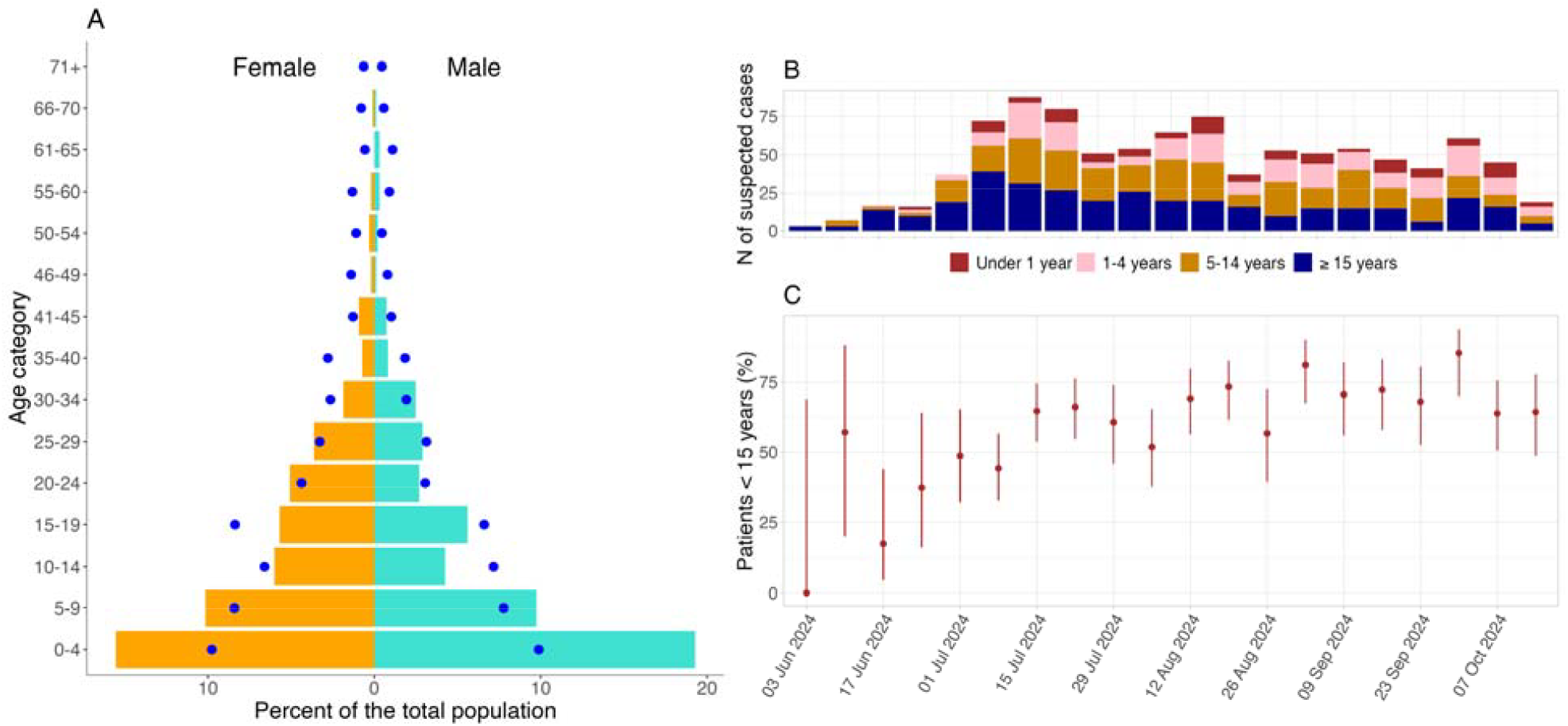
Description of the mpox epidemic in Uvira. Panel A: Age-sex pyramid of suspected cases and the general population of Uvira, with the bars representing the proportion of men (turquoise) and women (orange) by age group within the study population. The blue dots denote the disaggregation by age and sex from the population estimates based on household composition data from four cross-sectional surveys conducted in Uvira between 2021-2024 (n = 10,604 household members). Panel B: Epidemic curve by age group and week of presentation at the Mpox Treatment Center (MTC). Panel C: Weekly proportion of children under 15 years of age presenting to the MTC.

### Clinical case description, key populations and deaths

While all suspected cases were meant to have a sample collected and tested, only a subset had test results available due to laboratory supply and logistical limitations. Of 973 suspected cases, 415 (42.7%) were tested by at least one assay, with an overall mpox PCR positivity of 77.6% (322/415) (Table 2, Table S3, Table S4). Among 49 positive samples tested with the TIB Molbiol^®^ qPCR clade-typing assay, all were confirmed as clade Ib MPXV virus. Patients with laboratory test results had similar socio-demographic characteristics as those without test results but had more symptoms and lesions (Table S3). Patients testing positive had socio-demographic and clinical characteristics comparable to those with negative results (Table S4).

**Table 2.** Description of clinical characteristics of suspected cases on admission, by age. ^*^ Child’s breastfeeding status was only asked for those aged ≤ 2 years. ^**^ Questions on breastfeeding and pregnancy were only asked to female patients aged 15-49 years.

| Characteristic | Overall<br>(n = 973) | Under 15 years<br>(n = 620) | $\geq 15$ years<br>(n= 353) | p-value |
| --- | --- | --- | --- | --- |
| Hospitalized | 431/860 (50.1%) | 273/554 (49.3%) | 158/306 (51.6%) | 0.508 |
| Self-reported fever | 648/836 (77.5%) | 426/536 (79.5%) | 222/300 (74.0%) | 0.069 |
| Lesions on palms and soles | 320/872 (36.7%) | 200/561 (35.7%) | 120/311 (38.6%) | 0.389 |
| Cervical adenopathy | 550/870 (63.2%) | 400/559 (71.6%) | 150/311 (48.2%) | <0.001 |
| Inguinal adenopathy | 664/872 (76.1%) | 422/561 (75.2%) | 242/311 (77.8%) | 0.390 |
| Axillary adenopathy | 288/867 (33.2%) | 214/557 (38.4%) | 74/310 (23.9%) | <0.001 |
| Generalized adenopathy | 205/840 (24.4%) | 160/538 (29.7%) | 45/302 (14.9%) | <0.001 |
| Eye redness | 113/861 (13.1%) | 69/554 (12.5%) | 44/307 (14.3%) | 0.435 |
| Eye itching | 74/513 (14.4%) | 33/209 (15.8%) | 41/304 (13.5%) | 0.466 |
| Light sensitivity | 84/489 (17.2%) | 36/193 (18.7%) | 48/296 (16.2%) | 0.485 |
| Oral lesions | 204/852 (23.9%) | 135/549 (24.6%) | 69/303 (22.8%) | 0.552 |
| Genital lesions | 413/860 (48.0%) | 217/554 (39.2%) | 196/306 (64.1%) | <0.001 |
| Anal lesions | 123/852 (14.4%) | 73/548 (13.3%) | 50/304 (16.4%) | 0.214 |
| Number of lesions |  |  |  | <0.001 |
| <25 lesions | 238/858 (27.7%) | 123/554 (22.2%) | 115/304 (37.8%) |  |
| 25-99 lesions | 429/858 (50.0%) | 289/554 (52.2%) | 140/304 (46.1%) |  |
| 100-250 lesions | 125/858 (14.6%) | 102/554 (18.4%) | 23/304 (7.57%) |  |
| >250 lesions | 66/858 (7.69%) | 40/554 (7.2%) | 26/304 (8.5%) |  |
| Pregnant* | 19/131 (14.5%) | NA | 19/131 (14.5%) | NA |
| Breastfeeding mother* | 27/120 (22.5%) | NA | 27/120 (22.5%) | NA |
| Breastfed child** | 128/166 (77.1%) | 128/166 (77.1%) | NA | NA |
| Nutritional status of children 3 to 59 months old |  |  |  | NA |
| Severe acute malnutrition | 10/181 (5.52%) | 10/181 (5.52%) | NA |  |
| Moderate acute malnutrition | 12/181 (6.63%) | 12/181 (6.63%) | NA |  |
| Normal | 159/181 (87.8%) | 159/181 (87.8%) | NA |  |
| Syphilis test result |  |  |  | 0.374 |
| Negative | 272/273 (99.6%) | 171/171 (100.0%) | 101/102 (99.0%) |  |
| Positive | 1/273 (0.37%) | 0/171 (0%) | 1/102 (0.98%) |  |
| HIV test result |  |  |  | 0.002 |
| Negative | 323/329 (98.2%) | 214/214 (100.0%) | 109/115 (94.8%) |  |
| Positive | 6/329 (1.8%) | 0/214 (0%) | 6/115 (5.2%) |  |
| Mpox confirmation (PCR) |  |  |  | 0.031 |
| Negative | 93/415 (22.4%) | 68/264 (25.8%) | 25/151 (16.6%) |  |
| Positive | 322/415 (77.6%) | 196/264 (74.2%) | 126/151 (83.4%) |  |

\*Child's breastfeeding status was only asked for those aged $\leq 2$ years. \*\*Questions on breastfeeding and pregnancy were only asked to female patients aged 15-49 years.

**Table 3.** Description of exposure location, relationships with suspected Mpox cases, and contact characteristics. All questions refer to the 3 weeks before symptom onset. Contact history with suspected mpox cases was available for 346 patients, some of whom reported exposure to multiple suspected cases or multiple locations.

| Characteristic | Overall<br>(n = 460) | Under 15 years<br>(n = 321) | ≥15 years<br>(n = 139) | p-value |
| --- | --- | --- | --- | --- |
| Sex |  |  |  | <0.001 |
| male | 239/460 (52.0%) | 185/321 (57.6%) | 54/139 (38.8%) |  |
| female | 221/460 (48.0%) | 136/321 (42.4%) | 85/139 (61.2%) |  |
| Number of contacts |  |  |  | 0.006 |
| multiple | 373/401 (93.0%) | 266/279 (95.3%) | 107/122 (87.7%) |  |
| once | 28/401 (6.98%) | 13/279 (4.66%) | 15/122 (12.3%) |  |
| Relationship with the suspected case |  |  |  |  |
| Child | 30/417 (7.19%) | 0/287 (0%) | 30/130 (23.1%) |  |
| Parent | 13/417 (3.12%) | 11/287 (3.83%) | 2/130 (1.54%) |  |
| Spouse | 11/417 (2.64%) | 0/287 (0%) | 11/130 (8.46%) |  |
| Sex partner | 8/417 (1.92%) | 0/287 (0%) | 8/130 (6.15%) |  |
| Another household member (e.g. sibling) | 197/417 (47.2%) | 161/287 (56.1%) | 36/130 (27.7%) |  |
| Another non-household relative, friend, colleague, neighbor | 152/417 (36.5%) | 112/287 (39.0%) | 40/130 (30.8%) |  |
| Health care occupational exposure | 6/417 (1.4%) | 3/287 (1.1%) | 3/130 (2.3%) |  |
| Duration of contact |  |  |  | 0.100 |
| < 15 minutes | 8/29 (27.6%) | 2/17 (11.8%) | 6/12 (50.0%) |  |
| 15–60 minutes | 9/29 (31.0%) | 7/17 (41.2%) | 2/12 (16.7%) |  |
| > 60 minutes | 12/29 (41.4%) | 8/17 (47.1%) | 4/12 (33.3%) |  |
| Contact happened in Uvira | 423/439 (96.4%) | 298/304 (98.0%) | 125/135 (92.6%) | 0.010 |
| Reported contact person was hospitalized | 191/231 (82.7%) | 145/169 (85.8%) | 46/62 (74.2%) | 0.039 |
| Exposure at restaurants, bars, hotels, night clubs | 108/439 (24.6%) | 75/304 (24.7%) | 33/135 (24.4%) | 0.959 |
| Household exposure | 298/439 (67.9%) | 208/304 (68.4%) | 90/135 (66.7%) | 0.716 |
| Exposure at workplace | 7/439 (1.59%) | 0/304 (0%) | 7/135 (5.19%) | <0.001 |
| Exposure at school | 3/439 (0.68%) | 3/304 (0.99%) | 0/135 (0%) | 0.556 |
| Exposure at sport place | 1/439 (0.23%) | 1/304 (0.33%) | 0/135 (0%) | >0.999 |
| Exposure at health facility | 14/439 (3.19%) | 6/304 (1.97%) | 8/135 (5.93%) | 0.039 |

Severe to very severe mpox (≥100 lesions) was observed in 22.3% (n=191/858) of all cases, with a higher proportion in children under 15 years of age (25.6%, n = 142/554) compared with older individuals (16.1%, 49/304, p <0.001) (Table 2). A similar trend was seen in confirmed cases, where 31.6% (61/193) of children under 15 had of severe to very severe, versus 17.1% (20/117) of individuals aged ≥15 years.

(Table S5). Genital lesions were present in 48.0% (413/860) of all cases, significantly more frequent in individuals aged ≥15 years (64.1%, 196/306) than in younger patients (39.2%, 217/554; p < 0.001). This pattern was even more marked in confirmed cases, where 75.2% (88/117) of patients aged ≥15 years had genital lesions compared to 45.6% (88/193) of younger patients (p < 0.001), resulting in an overall incidence of 56.8% (176/310) among confirmed cases (Table S5). Anal lesions were reported in 14.4% (123/852) of suspected cases and 17.6% (54/306) of confirmed cases, with no significant differences by age or sex (Table 2 and Table S5). Overall, lymphadenopathy, particularly cervical, axillary, and generalized, was more common in patients under 15 years than in older individuals, in both suspected and confirmed cases.

Among 329 patients who consented to HIV testing, six (1.8%) had a positive test result, all of whom were aged ≥15 years (5.2% age-group positivity, 6/115), and with no significant difference by sex (Table 2 and Supplementary Table 2). One (0.4%) case of syphilis was detected among the 272 tested cases. Of 131 female patients of reproductive age (15–49 years), 14.5% (n=19) self-reported as pregnant, with 36.8% (7/19) in their third trimester (Table S5), and 3 (15.8%) presenting with severe to very severe (≥100 lesions) suspected mpox. By mid-December 2024, at the time of data analysis, follow-up data were available for 8 of the 19 pregnant patients, each with one follow-up visit. Among these, three had a positive PCR test, 1 a negative PCR and 4 with no lab results. They delivered nine live newborns (including one set of twins), with no complications or lesions reported by mothers. Two additional pregnant women followed up were still pregnant with no complication reported. Among children under five with MUAC measurements, 12.1% (22/181) had acute malnutrition, including 10 (5.5%) children with severe acute malnutrition and 12 (6.6%) with moderate malnutrition (Table 2).

Seven cases at the MTC died during the study period, with ages ranging between 13 days and 32 years (Supplementary Table S5). The overall case fatality ratio was 0.7% among all suspected cases. Time from hospitalization to death ranged from three to 29 days. Six of the seven patients who died had PCR results, of whom five tested positive, one tested negative, and one had no result. Five deaths occurred in infants under one year of age, including three infants under one month-old, yielding a case fatality ratio of 4% (5/127) in this age group. The two additional deaths occurred in immunocompromised adults with HIV, one of whom presented with disseminated necrotizing lesions (described elsewhere [30]), and the other with coexisting diabetes. Among cases that tested positive for HIV, the case fatality ratio was 33.3% (2/6). Detailed clinical descriptions of the deaths are provided in Table S6.

### Mpox exposures

Exposure data were collected from 743 patients for the three weeks preceding symptom onset. Of these, 46.6% (n=346) reported direct contact with a suspected mpox case in the three weeks prior to symptom onset, including 79.8% (276/346) exposed to a single case and 20.2% (70/346) exposed to multiple suspected cases, with no significant difference by sex (Table 1). Ninety (10.7%) patients reported travelling in the past three weeks, including 31 (34.4%) who traveled internationally, of whom 30 (96.8%) traveled to neighboring Burundi. Only 5.0% (38/776) of patients reported contact with animals in the three weeks prior to onset of symptoms, with most contacts involving domestic animals (97.4%, 38/39). No contact with rodents was reported. Health care occupational exposures accounted for 1.4% (6/417) of all exposures.

Overall, 70.5% (202/286) of cases aged ≥15 years reported having at least one sexual partner in the three weeks prior to symptom onset, and 17.1% (49/286) reported multiple sexual partners. Only 6% (19/318) of those who reported exposure to a suspected case, and for whom the relationship with the suspected case was known, identified sexual contact as the mode of exposure (Table 1).

Reported exposures to suspected mpox were primarily from within case households (67.9%, 298/439). Among them, 62.7% (187/298) were exposed to multiple suspected cases (Table 2). 3.2% of cases (14/439) reported exposure within a healthcare facility. Other exposures were reported in restaurants, bars, hotels, and nightclubs (24.6%, 108/439).

## Discussion

The clade Ib mpox outbreak in Uvira primarily affected children under 15 years (63.7%), with limited reported sexual transmission. Occupational healthcare and workplace exposures were rare (1.4% and 1.6%, respectively). The overall case fatality ratio was 0.7%, but immunocompromised patients and infants under one year faced notably high mortality ratio.

Several factors likely contributed to the high incidence of mpox clade Ib among children: (i) cryptic transmission, (ii) prior smallpox immunity in older individuals, and (iii) age-related differences in MPXV susceptibility, exacerbated by child malnutrition. Remarkably, the age distribution of clade Ib cases in Uvira mirrors patterns from historical clade Ia outbreaks.[2,31] However, despite higher mortality among young children, the CFR for clade Ib in infants younger than one year is less than half the 10% CFR reported in pediatric populations during clade Ia outbreaks.[5,32] This lower CFR may reflect the reduced virulence of clade Ib, possibly due to OPG032 gene deletion, which encodes the complement control protein, a key virulence factor.[33,34] Additionally, in remote and hard-to-reach areas where most historical clade Ia outbreaks occurred, surveillance bias may have led to an overestimation of CFR, as only the most severe cases would seek care, with milder cases likely remaining at home, thus biasing the denominator downward. Finally, improved and facilitated healthcare access in Uvira also likely reduced mortality. Unlike previous remote outbreaks in remote and hard-to-reach areas, this one received international support, with non-governmental organizations and the Ministry of Health providing free treatment, food, and hygiene kits.

This outbreak reflects a shift from sexual to non-sexual transmission, with overcrowded housing likely accelerating the spread. Thus, in Uvira where the median household size is eight and a median of four people sleep in each room, the WHO’s home isolation guidance is impractical, a situation common to many similar settings. While isolation at the MTC can reduce household spread, it imposes indirect costs on families and strains on the health system. These challenges suggest the potential for household-level interventions, such as “ring” vaccination,[35] or case-area targeted interventions (CATIs) used in cholera outbreaks, to curb intra-household transmission.[36]

The PCR positivity rate in this study was higher among individuals aged ≥15 years. This trend was also observed in Kamituga, where positivity was significantly higher in those ≥15 compared to <15 years (84.5% vs. 66.2%, p < 0.001).[13] Possible explanations include a likely higher burden of non-mpox rash illnesses (e.g., measles) in children attending MTCs, or differences in lesion type and location. Genital lesions, shown to be more common in older age groups, may carry higher viral loads;[37] if they are also more likely to be sampled (though we lack data on this), this could contribute to the higher PCR positivity rates observed in adults. Finally, samples were collected at random, mainly based on availability of materials. While we have no evidence of bias in sample collection, we cannot exclude the possibility that certain populations were preferentially sampled. Further investigations are needed to better understand the driver of differential mpox PCR positivity by age.

Our findings underscore the urgent need for integrated mpox programs targeting key vulnerable populations. HIV prevalence among suspected mpox cases was nearly three times higher than the estimated national average of 0.7%,[38] and nine times higher than the 0.2% prevalence observed among voluntary blood donors at Uvira Hospital in 2024. Alarmingly, all adult mpox-related deaths (n=2) occurred in HIV-positive individuals with severe necrotizing disease. Limited access to CD4 monitoring and opportunistic infection management complicates treatment, highlighting the importance of integrated mpox-HIV care models and vaccine safety studies for immunocompromised individuals. However, HIV prevalence estimates from the MTC should be interpreted with caution, as not all patients were tested, and selection bias cannot be excluded. Among children, undernutrition is the leading cause of acquired immune deficiency, and is especially concerning in the DRC where nearly half of children under five are stunted.[39] Our study found acute malnutrition rates (12.1%) among suspected mpox cases nearly double the 2023–2024 South Kivu province average of 6.7%.[39] This figure is likely an underestimate, as we relied solely on mid-upper arm circumference (MUAC) measurements without weight-for-height z-scores-scores.[40] Lastly, pregnant and breastfeeding women, who represented 14.5% and 22.5% of women of reproductive age at the Uvira MTC, respectively, also require specific attention given the height CFR of mpox clade Ib in neonates and emerging evidence suggesting potential mother-to-child transmission, including through breast milk.[41] Thus, our findings support WHO recommendations advocating for the vaccination of pregnant and lactating women,[42,43] and emphasize the need for greater investment in understanding the interplay between mpox clade Ib, age, malnutrition, HIV, and pregnancy. Addressing these gaps requires urgent attention, in-depth investigations, and targeted interventions that extend beyond current emergency responses.

A stark difference was however observed between our findings, which indicated no mpox-related adverse pregnancy outcomes, and those from Kamituga, a major mining hub where 57.8% (8/14) of pregnant women with mpox Clade Ib reported abortion, and mpox DNA was detected in one placenta. [41] Conflicting evidence on mother-to-child transmission of clade II mpox further highlights existing knowledge gaps.[5,44] Differences in environmental exposures, mpox severity, mode of acquisition, and co-infections, such as HIV or congenital co-infections may contribute to the observed disparities. In South Kivu, birth defect rates were nearly four times higher in mining areas like Mwenga, near Kamituga, even before the mpox outbreak.[45] Research from Lubumbashi, over 1,000 km south, linked paternal mining exposure, prenatal metal levels, and poor maternal nutrition to malformations in mining sites.[46] Additionally, Kamituga’s high commercial sex activity raises concerns about co-infections that may exacerbate pregnancy complications. Only one syphilis case was detected among 276 patients in Uvira, with no comparable data from Kamituga. Larger studies are needed to clarify the potential causal relationship between mpox clade Ib infection and pregnancy outcomes.

This study has several limitations. It relied on facility-based surveillance from MTC due to the lack of data on cases in private facilities or the community. The clinical case definition used in the MTC was broad, and the low proportion of cases tested may have led to misclassification. However, the high positivity rate among those tested provides some assurance that case definition was generally adequate. Some mpox cases could have not been managed in MTC, limiting their recording in our dataset.

Additionally, many patients were admitted to the MTC primarily for isolation, sometimes at their own request, rather than for inpatient care, limiting the interpretability of the significance of hospitalization status in our data set, particularly in the outbreak’s early months. The dataset also lacked precision regarding lesion distribution. For example, due to the structure of the questionnaire used in routine surveillance, we were unable to distinguish patients with isolated genital lesions from those with genital lesions in addition to lesions elsewhere on the body, limiting our ability to assess the role of sexual transmission in Uvira compared to other settings like Kamituga.

Our reliance on the GeneXpert®L Xpert Mpox assay posed several challenges. While the high positivity rate supports the clinical case definition used, this test detects MPXV Clade II and non-variola orthopoxviruses but cannot directly confirm Clade I. In our context, Clade I was inferred when results were positive for orthopoxvirus and negative for Clade II. Although this approach aligns with WHO guidance and is common in DRC, its performance, especially for Clade Ib, remains poorly characterized. However, a clade-typed subsample from Uvira confirmed all tested cases as Clade Ib, providing some reassurance.

Operational constraints further limited diagnostic accuracy. These included the absence of on-site testing, delays in sample transport and processing which may have affected specimen quality, GeneXpert stockouts, competing demands from tuberculosis testing, and communication gaps with laboratories.

Testing for other common causes of skin lesions (e.g., varicella-zoster, measles, herpes simplex virus) was not routinely performed, making it difficult to determine the etiology of mpox-negative cases.

Misclassification, whether from clinical diagnosis or imperfect testing, not only affects burden estimation but also raises infection control concerns. Patients admitted to MTCs based on clinical suspicion alone may have been exposed to, or may have exposed others to, infectious diseases. For this reason, we included both confirmed and unconfirmed cases to reflect the real-world complexity of mpox management and to highlight the urgent need for decentralized diagnostics, dedicated platforms, and stronger laboratory networks in outbreak-prone settings.

In conclusion, the Uvira outbreak signals a major shift in mpox transmission, with household-driven spread disproportionately affecting children. This pattern, distinct from the clade II outbreaks in men who have sex with men and early clade Ib cases in Kamituga linked to heterosexual transmission, presents a global risk. Response efforts in DRC, however, have been shaped by the epidemiology in Kamituga with vaccination targeting key adult populations. However, this strategy may be less effective in areas like Uvira, where transmission has already entrenched, particularly among young children. Surveillance suggests similar patterns of non-sexual household transmission may be occurring throughout much of South Kivu, emphasizing the need for broader interventions. Sustained child-centered transmission could fuel widespread epidemics, particularly in high-birth rate, resource-limited settings where residual protection from smallpox vaccination efforts is largely gone. Urgent global action including accelerating pediatric vaccine development, expanding therapeutic access, and implementing household-level interventions, is essential to contain clade Ib and mitigate its long-term health impacts on individuals and populations.

## Data Availability

All data produced in the present work are contained in the manuscript

## Patient Consent Information

The data used in this study were collected with the Uvira health zone team for public health surveillance. Ethical approvals were obtained from the Institutional Review Boards of the Johns Hopkins Bloomberg School of Public Health (reference number IRB00030442) and the Université Catholique de Bukavu (UCB/CIES/NC/019/2024) to use those data.

## References

1. Jezek Z, Grab B, Szczeniowski MV, Paluku KM, Mutombo M. Human monkeypox: secondary attack rates. Bull World Health Organ. 1988;66(4):465–70.

2. Breman JG, Kalisa-Ruti Steniowski MV, Zanotto E, Gromyko AI, Arita I. Human monkeypox, 1970-79. Bull World Health Organ. 1980;58(2):165–82.

3. Bangwen E, Diavita R, De Vos E, Hasivirwe Vakaniaki E, Nundu SS, Mutombo A, et al. Mpox in the democratic republic of Congo: Analysis of national epidemiological and laboratory surveillance data, 2010 - 2023 [Internet]. 2024. Available from: 10.2139/ssrn.4954317

4. Bunge EM, Hoet B, Chen L, Lienert F, Weidenthaler H, Baer LR, et al. The changing epidemiology of human monkeypox-A potential threat? A systematic review. PLoS Negl Trop Dis. 2022 Feb;16(2):e0010141.

5. Sanchez Clemente N, Coles C, Paixao ES, Brickley EB, Whittaker E, Alfven T, et al. Paediatric, maternal, and congenital mpox: a systematic review and meta-analysis. Lancet Glob Health. 2024 Apr;12(4):e572–88.

6. Gessain A, Nakoune E, Yazdanpanah Y. Monkeypox. N Engl J Med. 2022 Nov 10;387(19):1783– 93.

7. Mitjà O, Ogoina D, Titanji BK, Galvan C, Muyembe JJ, Marks M, et al. Monkeypox. Lancet. 2023 Jan 7;401(10370):60–74.

8. Vakaniaki EH, Kacita C, Kinganda-Lusamaki E, O’Toole Á, Wawina-Bokalanga T, Mukadi-Bamuleka D, et al. Sustained human outbreak of a new MPXV clade I lineage in eastern Democratic Republic of the Congo. Nature Medicine. 2024 Jun 13;30(10):2791–5.

9. Mukadi-Bamuleka D, Kinganda-Lusamaki E, Mulopo-Mukanya N, Amuri-Aziza A, O’Toole Á, Modadra-Madakpa B, et al. First imported Cases of MPXV Clade Ib in Goma, Democratic Republic of the Congo: Implications for Global Surveillance and Transmission Dynamics. medRxiv [Internet]. 2024 Sep 16; Available from: 10.1101/2024.09.12.24313188

10. Nizigiyimana A, Ndikumwenayo F, Houben S, Manirakiza M, van Lettow M, Liesenborghs L, et al. Epidemiological analysis of confirmed mpox cases, Burundi, 3 July to 9 September 2024. Euro Surveill [Internet]. 2024 Oct;29(42). Available from: 10.2807/1560-7917.ES.2024.29.42.2400647

11. WHO. WHO Director-General declares mpox outbreak a public health emergency of international concern [Internet]. WHO; 2024 [cited 2024 Dec 11]. Available from: https://www.who.int/news/item/14-08-2024-who-director-general-declares-mpox-outbreak-a-public-health-emergency-of-international-concern

12. Kibungu EM, Vakaniaki EH, Kinganda-Lusamaki E, Kalonji-Mukendi T, Pukuta E, Hoff NA, et al. Clade I-Associated Mpox Cases Associated with Sexual Contact, the Democratic Republic of the Congo. Emerg Infect Dis. 2024 Jan;30(1):172–6.

13. Brosius I, Vakaniaki EH, Mukari G, Munganga P, Tshomba JC, De Vos E, et al. Epidemiological and clinical features of mpox during the clade Ib outbreak in South Kivu, Democratic Republic of the Congo: a prospective cohort study. Lancet [Internet]. 2025 Jan; Available from: 10.1016/S0140-6736(25)00047-9

14. Masirika LM, Nieuwenhuijse DF, Ndishimye P, Udahemuka JC, Steeven BK, Gisèle NB, et al. Mapping the distribution and describing the first cases from an ongoing outbreak of a New Strain of mpox in South Kivu, Eastern Democratic Republic of Congo between September 2023 to April 2024 [Internet]. medRxiv. 2024 [cited 2024 Dec 2]. p. 2024.05.10.24307057. Available from: https://www.medrxiv.org/content/10.1101/2024.05.10.24307057v1.abstract

15. WHO. Mpox - Democratic Republic of the Congo. 2024 Jun 14 [cited 2024 Dec 10]; Available from: https://www.who.int/emergencies/disease-outbreak-news/item/2024-DON522

16. World Health Organization. Mpox in the WHO African region [Internet]. World Health Organization; 2024 Nov. Report No.: Weekly Regional Situation Report #12. Available from: https://iris.who.int/bitstream/handle/10665/379531/AFRO-Mpox%20bulletin-%2010%20November%202024.pdf

17. Koyuncu A, Bugeme PM, Hulse JD, Hutchins C, Xu H, Gallandat K, et al. Challenges with Achieving and Maintaining High Oral Cholera Vaccine Coverage in Uvira, The Democratic Republic of the Congo: serial cross-sectional representative surveys [Internet]. 2024. Available from: 10.31219/osf.io/fgq6e

18. De Keyzer ELR, Masilya Mulungula P, Alunga Lufungula G, Amisi Manala C, Andema Muniali A, Bashengezi Cibuhira P, et al. Local perceptions on the state of the pelagic fisheries and fisheries management in Uvira, Lake Tanganyika, DR Congo. J Great Lakes Res. 2020 Dec 1;46(6):1740–53.

19. UNICEF. Democratic Republic of the Congo Humanitarian Situation Report No. 1, 30 June 2024 [Internet]. UNICEF; 2024 Aug [cited 2025 Jun 16]. Report No.: 1. Available from: https://www.unicef.org/media/160561/file/DRC-Humanitarian-SitRep-30-June-2024.pdf

20. Mugisho GM, Swedi DP, Enock PM, Kalembu MI, Lukeba FN, Ngezirabona SV. Urban anthropization: community vulnerability and resilience to flood hazards in eastern Democratic Republic of Congo. Environ Res Commun. 2024 Feb 1;6(2):025003.

21. World Health Organization. Surveillance, case investigation and contact tracing for mpox : Interim guidance, 27 November 2024 [Internet]. World Health Organization; 2024 Nov [cited 2025 Jan 11]. Available from: https://pesquisa.bvsalud.org/portal/resource/pt/who-376306

22. World Health Organization and UNICEF. WHO child growth standards and the identification of severe acute malnutrition in infants and children A Joint Statement by the World Health Organization and the United Nations Children’s Fund [Internet]. World Health Organization and UNICEF; 2009. Available from: https://iris.who.int/bitstream/handle/10665/44129/9789241598163_eng.pdf

23. TIB Molbiol. LightMix Modular Monkeypox Virus 580 [Internet]. TIB Molbiol; 2022 [cited 2025 Jun 16]. Available from: https://assets.cwp.roche.com/f/94122/7eb8b239c4/mdx_58-0550-96_monkeypox_v220929_09798471001.pdf

24. Harshani HBC, Liyanage GA, Ruwan DVRG, Samaraweera UKIU, Abeynayake JI. Evaluation of the diagnostic performance of a commercial molecular assay for the screening of suspected monkeypox cases in Sri Lanka. Infect Med (Beijing). 2023 Jun;2(2):136–42.

25. Li D, Wilkins K, McCollum AM, Osadebe L, Kabamba J, Nguete B, et al. Evaluation of the GeneXpert for human Monkeypox diagnosis. Am J Trop Med Hyg. 2017 Feb 8;96(2):405–10.

26. Cepheid. Xpert® Mpox [Internet]. Cepheid; 2023 Feb. Available from: https://www.fda.gov/media/165322/download

27. Clinical Aspects of Monkey Pox in DR Congo : Monkeypox epidemiology, surveillance, and laboratory capacity in DRC.

28. Myatt M, Guevarra E. Package “zscorer” [Internet]. 2019 [cited 2025 Feb 15]. Available from: https://cran.r-project.org/web/packages/zscorer/zscorer.pdf

29. Arm circumference-for-age [Internet]. [cited 2025 Feb 15]. Available from: https://www.who.int/tools/child-growth-standards/standards/arm-circumference-for-age

30. Bugeme PM, Bugale PK, Mashauri JZ, Kapalata AI, Bugwaja L, Trust Faraja Mukika, et al. Disseminated mpox in an immunocompromised patient in DR Congo: a call for a shift from disease-focused to person-centred outbreak response. Lancet Infect Dis [Internet]. 2025 Jan; Available from: 10.1016/s1473-3099(25)00009-x

31. Jezek Z, Marennikova SS, Mutumbo M, Nakano JH, Paluku KM, Szczeniowski M. Human monkeypox: a study of 2,510 contacts of 214 patients. J Infect Dis. 1986 Oct;154(4):551–5.

32. Jezek Z, Szczeniowski M, Paluku KM, Mutombo M. Human monkeypox: clinical features of 282 patients. J Infect Dis. 1987 Aug;156(2):293–8.

33. Sganzerla Martinez G, Kumar A, Kinganda Lusamaki E, Dutt M, Wawina-Bokalanga T, Toloue Ostadgavahi A, et al. Monkeypox virus pangenomics reveals determinants of clade Ib [Internet]. medRxiv. 2024. Available from: 10.1101/2024.10.31.24315917

34. Hudson PN, Self J, Weiss S, Braden Z, Xiao Y, Girgis NM, et al. Elucidating the role of the complement control protein in monkeypox pathogenicity. PLoS One. 2012 Apr 9;7(4):e35086.

35. Muyembe JJ, Pan H, Peto R, Diallo A, Touré A, Mbala-Kingebene P, et al. Ebola outbreak response in the DRC with rVSV-ZEBOV-GP ring vaccination. N Engl J Med. 2024 Dec 19;391(24):2327–36.

36. Ratnayake R, Finger F, Azman AS, Lantagne D, Funk S, Edmunds WJ, et al. Highly targeted spatiotemporal interventions against cholera epidemics, 2000-19: a scoping review. Lancet Infect Dis. 2021 Mar;21(3):e37–48.

37. Palich R, Burrel S, Monsel G, Nouchi A, Bleibtreu A, Seang S, et al. Viral loads in clinical samples of men with monkeypox virus infection: a French case series. Lancet Infect Dis. 2023 Jan;23(1):74– 80.

38. UNAIDS. Country factsheets : Democratic Republic of the Congo, 2023 [Internet]. Democratic Republic of the Congo - UNAIDS. Available from: https://www.unaids.org/en/regionscountries/countries/democraticrepublicofthecongo

39. RDC-Institut National de la Statistique, École de Santé Publique de Kinshasa et ICF. 2024. RDC, Enquête Démographique et de Santé 2023–24 : Rapport des indicateurs clés. Kinshasa, RDC et Rockville, Maryland, USA : ICF. Available from: https://dhsprogram.com/pubs/pdf/PR156/PR156.pdf

40. Grellety E, Golden MH. Weight-for-height and mid-upper-arm circumference should be used independently to diagnose acute malnutrition: policy implications. BMC Nutr [Internet]. 2016 Dec;2(1). Available from: 10.1186/s40795-016-0049-7

41. Masirika LM, Udahemuka JC, Schuele L, Nieuwenhuijse DF, Ndishimye P, Boter M, et al. Epidemiological and genomic evolution of the ongoing outbreak of clade Ib mpox virus in the eastern Democratic Republic of the Congo. Nat Med [Internet]. 2025 Feb 11; Available from: 10.1038/s41591-025-03582-1

42. Hombach J, Lewis R, van Holten J, Scott JA, Neuzil KM. Breastfeeding mothers in DR Congo should have access to the mpox vaccine. Lancet Glob Health. 2024 Dec 1;12(12):e1932.

43. Health Organization (WHO) W. Smallpox and Mpox (Orthopoxviruses): WHO Position Paper. WHO: Geneva, Switzerland. 2024;99:429–56.

44. Marcela MR, Marcela D, Yazmín RP, Paula AM, Carlos FM, Hector RM, et al. Monkeypox virus infection in pregnancy: description of two cases reported to the Colombian National Institute of Health. APMIS. 2025 Jan;133(1):e13488.

45. Cikomola FG, Bisimwa AW, Nyalundja AD, Barthélemy EJ, Matabaro BS, Mukamba FM, et al. Prevalence and risk of occurrence of visible birth defects in mining areas in South Kivu: A hospital-based cross-sectional study. PLoS One. 2024 Oct 7;19(10):e0309004.

46. Van Brusselen D, Kayembe-Kitenge T, Mbuyi-Musanzayi S, Lubala Kasole T, Kabamba Ngombe L, Musa Obadia P, et al. Metal mining and birth defects: a case-control study in Lubumbashi, Democratic Republic of the Congo. Lancet Planet Health. 2020 Apr;4(4):e158–67.

