## Supplementary file for "Community transmission of Mpox clade Ib not driven through sexual exposures in Uvira, eastern Democratic Republic of Congo, June – October 2024" (from another version of this preprint: version 1, 2025-02-26)

Supplementary document

Patrick Musole Bugeme^1, 2, 3^, Patrick Kazuba Bugale^3^, Trust Faraja Mukika^3^, Megan O’Driscoll^4^,  Javier Perez-Saez^4^, Levi Bugwaja^1,2^, Salomon Mashupe Shangula^3^, Willy Kasi^5^, Justin Bengehya^5^, Stephanie Ngai^6^, Antonio Isidro Carrion Martin^7^, Jules Jackson^1^, Patrick Katoto^2^, Esto Bahizire^2,8^,  Noella Mulopo-Mukanya^9^, Justin Lessler^1,10,11^ Jackie Knee^12^, Pauline Vetter^4^,  Elizabeth C. Lee^1^, Daniel Mukadi-Bamuleka^10,13,14^, Andrew S Azman^1,4,15^, Espoir Bwenge Malembaka^1,2^*

*^1^Department of Epidemiology, Johns Hopkins University, Baltimore, MD 21205, USA*

*^2^Center for Tropical Diseases and Global Health (CTDGH), Université Catholique de Bukavu, Bukavu, Democratic Republic of the Congo*

*^3^Hôpital Général de Référence d’Uvira, Uvira Health Zone, Uvira, Democratic Republic of the Congo*

*^4^Geneva Centre for Emerging Viral Diseases, Geneva University Hospitals, Geneva, Switzerland,*

*^5^Division Provinciale de la Santé Publique du Sud-Kivu, Bukavu, Democratic Republic of the Congo*

*^6^Médecins Sans Frontières, Uvira, The Democratic Republic of the Congo*

*^7^Médecins Sans Frontières, London, United Kingdom*

*^8^Centre de Recherche en Sciences Naturelles de Lwiro, Bukavu, DR Congo*

*^9^Rodolphe Merieux INRB-Goma Laboratory, Goma, Democratic Republic of Congo*

*^10^University of North Carolina Population Center, University of North Carolina at Chapel Hill, Chapel Hill, NC, USA*

*^11^Department of Epidemiology, Gillings School of Global Public Health, University of North Carolina at Chapel Hill, Chapel Hill, NC, USA*

*^12^Department of Disease Control, London School of Hygiene & Tropical Medicine, London, UK*

*^13^Department of Virology, Institut National de Recherche Biomédicale (INRB), Kinshasa, Democratic Republic of the Congo*

*^14^Service de Microbiologie, Département de Biologie Médicale, Cliniques Universitaires de Kinshasa, Université de Kinshasa, Kinshasa, Democratic Republic of the Congo*

*^15^Division of Tropical and Humanitarian Medicine, Geneva University Hospitals (HUG), Geneva, Switzerland*

*Correspondence to:
Espoir Bwenge Malembaka

**Content**

### 1. **Mpox case definition and description of patients**

The DRC’s Ministry of Health, through the Programme National de Lutte contre le Monkeypox et les Fièvres Hémorragiques Virales (PNLMPX-FHV), considers a suspect mpox case as 1) anyone with a sudden high fever followed by a vesiculo-pustular rash predominantly on the face and present on the palms of the hands and soles of the feet; 2) or presence of at least 5 smallpox-type scars, 3) or anyone with fever > 38.3°C (101 F), severe headache, lymphadenopathy, back pain, myalgia, and severe asthenia, followed one to three days later by a progressive rash that often begins on the face (the densest) and then spreads elsewhere on the body, including the soles of the feet and palms of the hands. The community case definition of mpox is any person presenting with fever and skin rash. At the Uvira MTC, after the outbreak was confirmed, anyone presenting with a skin rash or lesions was considered a suspected mpox case and eligible for this study (Table S1).

**Table S1. Socio-demographics and mpox exposure in Uvira by sex**

| Characteristic | Overall  (n = 973) | Female  (n = 491) | Male  (n= 482) | p-value |
| --- | --- | --- | --- | --- |
| Age in years, median (IQR) | 9 (3.0, 20.0) | 10 (4.0, 21.0) | 8 (3.0, 18.0) | 0.051 |
| Age group |  |  |  | 0.052 |
| Under 1 year | 127/973 (13.1%) | 63/491 (12.8%) | 64/482 (13.3%) |  |
| 1-4 years | 217/973 (22.3%) | 91/491 (18.5%) | 126/482 (26.1%) |  |
| 5-14 years | 291/973 (29.9%) | 156/491 (31.8%) | 135/482 (28.0%) |  |
| 15-44 years | 318/973 (32.7%) | 172/491 (35.0%) | 146/482 (30.3%) |  |
| ≥ 45 years | 20/973 (2.06%) | 9/491 (1.83%) | 11/482 (2.28%) |  |
| Household size | 8 (5.0, 10.0) | 8 (5.0, 10.0) | 8 (5.0, 10.0) | 0.466 |
| Number of children under 5 in the household |  |  |  | 0.578 |
| One to two | 404/632 (63.9%) | 204/320 (63.8%) | 200/312 (64.1%) |  |
| Three or more | 106/632 (16.8%) | 50/320 (15.6%) | 56/312 (17.9%) |  |
| Zero | 122/632 (19.3%) | 66/320 (20.6%) | 56/312 (17.9%) |  |
| Number of people per sleeping room in household, median (IQR)* |  |  |  | 0.358 |
| Median (Q1, Q3) | 4 (2.5, 5.0) | 4 (2.5, 5.0) | 4 (2.7, 5.5) |  |
| Student | 310/973 (31.9%) | 151/491 (30.8%) | 159/482 (33.0%) | 0.455 |
| Pre-school age children | 293/973 (30.1%) | 134/491 (27.3%) | 159/482 (33.0%) | 0.053 |
| Doing small business | 83/973 (8.53%) | 65/491 (13.2%) | 18/482 (3.73%) | <0.001 |
| Unemployed | 69/973 (7.09%) | 55/491 (11.2%) | 14/482 (2.90%) | <0.001 |
| Farmer | 18/973 (1.85%) | 13/491 (2.65%) | 5/482 (1.04%) | 0.062 |
| Sex worker | 12/973 (1.23%) | 12/491 (2.44%) | 0/482 (0%) | <0.001 |
| Educator/Teacher | 9/973 (0.92%) | 3/491 (0.61%) | 6/482 (1.24%) | 0.337 |
| Driver | 12/973 (1.23%) | 0/491 (0%) | 12/482 (2.49%) | <0.001 |
| Artisan (manual work) | 9/973 (0.92%) | 6/491 (1.22%) | 3/482 (0.62%) | 0.506 |
| Cyclist | 7/973 (0.72%) | 0/491 (0%) | 7/482 (1.45%) | 0.007 |
| Health care worker | 4/973 (0.41%) | 2/491 (0.41%) | 2/482 (0.41%) | >0.999 |
| Number of sexual partners in the past 3 weeks** |  |  |  | <0.001 |
| Zero | 84/286 (29.4%) | 38/149 (25.5%) | 46/137 (33.6%) |  |
| One | 153/286 (53.5%) | 96/149 (64.4%) | 57/137 (41.6%) |  |
| 2 to 5 | 42/286 (14.7%) | 9/149 (6.04%) | 33/137 (24.1%) |  |
| At least 6 | 7/286 (2.45%) | 6/149 (4.03%) | 1/137 (0.73%) |  |
| Had contact with a suspected mpox case*** | 346/743 (46.6%) | 174/381 (45.7%) | 172/362 (47.5%) | 0.614 |
| Number of contacts with a suspected mpox case*** |  |  |  | 0.087 |
| One | 276/346 (79.8%) | 146/174 (83.9%) | 130/172 (75.6%) |  |
| Two | 43/346 (12.4%) | 15/174 (8.62%) | 28/172 (16.3%) |  |
| At least 3 | 27/346 (7.80%) | 13/174 (7.47%) | 14/172 (8.14%) |  |
| Sexual exposure to a suspected mpox case** | 19/318 (5.97%) | 12/158 (7.59%) | 7/160 (4.38%) | 0.226 |
| Working in mining site | 4/973 (0.41%) | 1/491 (0.20%) | 3/482 (0.62%) | 0.370 |
| Had contact with animal | 39/776 (5.03%) | 17/395 (4.30%) | 22/381 (5.77%) | 0.349 |
| Type of animal contact |  |  |  | 0.436 |
| Domestic animals | 38/39 (97.4%) | 16/17 (94.1%) | 22/22 (100.0%) |  |
| Wild animals | 1/39 (2.56%) | 1/17 (5.88%) | 0/22 (0%) |  |
| Traveled outside Uvira*** | 90/845 (10.7%) | 40/426 (9.39%) | 50/419 (11.9%) | 0.231 |
| International travel*** |  |  |  | 0.419 |
| Burundi | 30/31 (96.8%) | 12/13 (92.3%) | 18/18 (100.0%) |  |
| Tanzania | 1/31 (3.23%) | 1/13 (7.69%) | 0/18 (0%) |  |

Data are median (IQR) or n/N (%), N representing the number of individuals with non-missing data. *Household characteristics data were not available for non-residents of Uvira city and Uvira residents for whom a household visit was not conducted mainly due lack of accurate residential address. **Sexual exposure was reported only for individuals aged ≥ 15 years. Occupation was a multiple-choice question. ***All questions about exposure and travel history referred to the three weeks preceding the onset of fever or skin rash.

**Table S2. Clinical description of suspected mpox cases sex**

| **Characteristic** | **Overall** (n = 973) | **Female** (n= 491) | **Male** (n= 482) | **p-value** |
| --- | --- | --- | --- | --- |
| Hospitalized | 431/860 (50.1%) | 225/434 (51.8%) | 206/426 (48.4%) | 0.307 |
| Self-reported fever | 648/836 (77.5%) | 326/425 (76.7%) | 322/411 (78.3%) | 0.570 |
| Lesions on palms and soles | 320/872 (36.7%) | 152/443 (34.3%) | 168/429 (39.2%) | 0.137 |
| Cervical adenopathy | 550/870 (63.2%) | 267/439 (60.8%) | 283/431 (65.7%) | 0.139 |
| Inguinal adenopathy | 664/872 (76.1%) | 325/442 (73.5%) | 339/430 (78.8%) | 0.066 |
| Axillary adenopathy | 288/867 (33.2%) | 137/439 (31.2%) | 151/428 (35.3%) | 0.203 |
| Generalized adenopathy | 205/840 (24.4%) | 101/425 (23.8%) | 104/415 (25.1%) | 0.662 |
| Eye redness | 113/861 (13.1%) | 58/437 (13.3%) | 55/424 (13.0%) | 0.896 |
| Eye itching | 74/513 (14.4%) | 42/271 (15.5%) | 32/242 (13.2%) | 0.464 |
| Light sensitivity | 84/489 (17.2%) | 46/261 (17.6%) | 38/228 (16.7%) | 0.779 |
| Oral lesions | 204/852 (23.9%) | 104/428 (24.3%) | 100/424 (23.6%) | 0.807 |
| Genital lesions | 413/860 (48.0%) | 193/433 (44.6%) | 220/427 (51.5%) | 0.041 |
| Anal lesions | 123/852 (14.4%) | 60/431 (13.9%) | 63/421 (15.0%) | 0.665 |
| Number of lesions |  |  |  | 0.266 |
| <25 lesions | 238/858 (27.7%) | 131/434 (30.2%) | 107/424 (25.2%) |  |
| 25-99 lesions | 429/858 (50.0%) | 212/434 (48.8%) | 217/424 (51.2%) |  |
| 100-250 lesions | 125/858 (14.6%) | 56/434 (12.9%) | 69/424 (16.3%) |  |
| >250 lesions | 66/858 (7.69%) | 35/434 (8.06%) | 31/424 (7.31%) |  |
| Pregnancy status* | 19/131 (14.5%) | 19/131 (14.5%) | - | - |
| Breastfeeding mother* | 27/120 (22.5%) | 27/120 (22.5%) | - | - |
| Breastfed child** | 128/166 (77.1%) | 67/80 (83.8%) | 61/86 (70.9%) | 0.049 |
| Nutritional status of children |  |  |  | 0.166 |
| Severe acute malnutrition | 10/181 (5.52%) | 2/83 (2.41%) | 8/98 (8.16%) |  |
| Moderate acute malnutrition | 12/181 (6.63%) | 7/83 (8.43%) | 5/98 (5.10%) |  |
| Normal | 159/181 (87.8%) | 74/83 (89.2%) | 85/98 (86.7%) |  |
| Syphilis test |  |  |  | >0.999 |
| Negative | 272/273 (99.6%) | 149/150 (99.3%) | 123/123 (100.0%) |  |
| Positive | 1/273 (0.37%) | 1/150 (0.67%) | 0/123 (0%) |  |
| HIV test |  |  |  | >0.999 |
| Negative | 323/329 (98.2%) | 174/177 (98.3%) | 149/152 (98.0%) |  |
| Positive | 6/329 (1.82%) | 3/177 (1.69%) | 3/152 (1.97%) |  |
| Mpox confirmation (PCR) |  |  |  | 0.992 |
| Negative | 93/415 (22.4%) | 45/201 (22.4%) | 48/214 (22.4%) |  |
| Positive | 322/415 (77.6%) | 156/201 (77.6%) | 166/214 (77.6%) |  |

*Questions on breastfeeding and pregnancy were only asked to female patients aged 15-49 years. **Child’s breastfeeding status was only asked for those aged ≤ 2 years.

2. **Mpox confirmation challenges in Uvira**

During the study period, on-site confirmation of mpox was unavailable. Skin lesion swab and/or oro-pharyngeal swab samples were stored at 2–8°C in a refrigerator at the Uvira Health Zone office, often for 1–3 weeks or longer, before being transported to the referral laboratory in Bukavu, 120 km away (a 4–6-hour drive). Testing at the Bukavu lab was conducted using GeneXpert®️platform (Xpert® Mpox, Cepheid, Sunnyvale, California, USA), on two 4-module machines originally designated for tuberculosis testing.

In the outbreak's early phases, all mpox samples from South Kivu, except those from the remote area of Kamituga, which had its own GeneXpert testing capacity, were sent to the Rodolphe Mérieux laboratory at the Institut National de Recherche Biomédicale (INRB) in Goma, North Kivu, for PCR confirmation. In June 2024, testing was decentralized. INRB Goma, in collaboration with the South Kivu health division and WHO, trained staff at the South Kivu laboratory and installed mpox GeneXpert modules on the existing tuberculosis machines. This enabled the South Kivu laboratory to handle samples from 33 of the province’s 34 health zones (including Uvira). However, logistical and operational challenges persisted, including frequent stockouts of sample collection materials and cartridges, logistical delays in sample transportation, and processing backlogs. During these periods, samples were rerouted to INRB Goma for testing.

The Bukavu laboratory, responsible for most of South Kivu, faced additional strain due to the dual use of machines for both mpox and tuberculosis testing, leading to breakdowns and delays in processing and result interpretation. Communication challenges between the reference labs in Bukavu or Goma and the Uvira MTC further compounded delays, with results sometimes arriving weeks after patients had been discharged. Additionally, used cartridges were stored at the Bukavu laboratory until transported to Uvira, the only site equipped with an incinerator capable of safely destroying them. Figure S1 below shows the weekly testing rate for samples from Uvira during the study period.

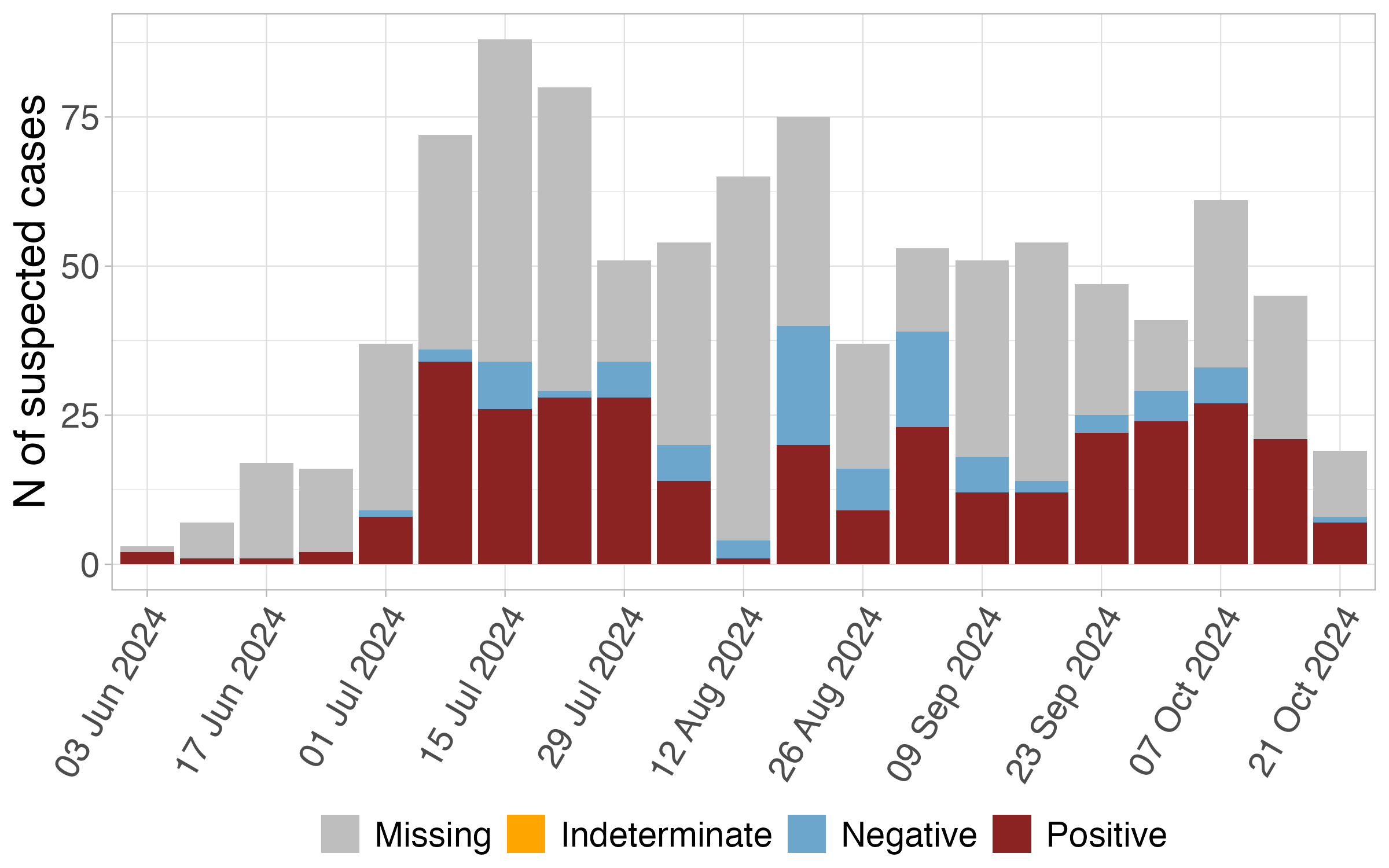

**Figure S1.** Epidemic curve of suspected mpox patients in Uvira by GeneXpert confirmation and week

A detailed comparison of tested and untested cases is presented in **Table S1**, while Table S 2 provides a comparison of positive and negative cases.

**Table S3. Sociodemographic and clinical characteristics of laboratory-tested suspected mpox cases**

| **Characteristic** | **Overall** (n = 973) | **Tested** (n = 415) | **Untested** (n= 558) | **p-value** |
| --- | --- | --- | --- | --- |
| Sex |  |  |  | 0.275 |
| Female | 491/973 (50.5%) | 201/415 (48.4%) | 290/558 (52.0%) |  |
| Male | 482/973 (49.5%) | 214/415 (51.6%) | 268/558 (48.0%) |  |
| Age in years, median (IQR) | 9 (3.0, 20.0) | 9 (2.8, 21.0) | 9 (4.0, 19.0) | 0.305 |
| Age group |  |  |  | 0.083 |
| Under 1 year | 127/973 (13.1%) | 65/415 (15.7%) | 62/558 (11.1%) |  |
| 1-4 years | 217/973 (22.3%) | 90/415 (21.7%) | 127/558 (22.8%) |  |
| 5-14 years | 291/973 (29.9%) | 114/415 (27.5%) | 177/558 (31.7%) |  |
| 15-44 years | 318/973 (32.7%) | 141/415 (34.0%) | 177/558 (31.7%) |  |
| ≥ 45 years | 20/973 (2.06%) | 5/415 (1.20%) | 15/558 (2.69%) |  |
| Hospitalized | 431/860 (50.1%) | 238/395 (60.3%) | 193/465 (41.5%) | <0.001 |
| Self-reported fever | 648/836 (77.5%) | 322/388 (83.0%) | 326/448 (72.8%) | <0.001 |
| Lesions on palms and soles | 320/872 (36.7%) | 172/397 (43.3%) | 148/475 (31.2%) | <0.001 |
| Cervical adenopathy | 550/870 (63.2%) | 289/399 (72.4%) | 261/471 (55.4%) | <0.001 |
| Inguinal adenopathy | 664/872 (76.1%) | 340/399 (85.2%) | 324/473 (68.5%) | <0.001 |
| Axillary adenopathy | 288/867 (33.2%) | 171/398 (43.0%) | 117/469 (24.9%) | <0.001 |
| Generalized adenopathy | 205/840 (24.4%) | 131/382 (34.3%) | 74/458 (16.2%) | <0.001 |
| Eye redness | 113/861 (13.1%) | 62/394 (15.7%) | 51/467 (10.9%) | 0.037 |
| Eye itching | 74/513 (14.4%) | 45/227 (19.8%) | 29/286 (10.1%) | 0.002 |
| Light sensitivity | 84/489 (17.2%) | 50/218 (22.9%) | 34/271 (12.5%) | 0.002 |
| Oral lesions | 204/852 (23.9%) | 125/392 (31.9%) | 79/460 (17.2%) | <0.001 |
| Genital lesions | 413/860 (48.0%) | 221/396 (55.8%) | 192/464 (41.4%) | <0.001 |
| Anal lesions | 123/852 (14.4%) | 62/392 (15.8%) | 61/460 (13.3%) | 0.290 |
| Number of lesions |  |  |  | 0.121 |
| <25 lesions | 238/858 (27.7%) | 99/395 (25.1%) | 139/463 (30.0%) |  |
| 25-99 lesions | 429/858 (50.0%) | 200/395 (50.6%) | 229/463 (49.5%) |  |
| 100-250 lesions | 125/858 (14.6%) | 68/395 (17.2%) | 57/463 (12.3%) |  |
| >250 lesions | 66/858 (7.69%) | 28/395 (7.09%) | 38/463 (8.21%) |  |
| Pregnancy status | 19/131 (14.5%) | 8/61 (13.1%) | 11/70 (15.7%) | 0.673 |
| Nutritional status of children |  |  |  | 0.484 |
| Severe acute malnutrition | 10/181 (5.52%) | 4/96 (4.17%) | 6/85 (7.06%) |  |
| Moderate acute malnutrition | 12/181 (6.63%) | 5/96 (5.21%) | 7/85 (8.24%) |  |
| Normal | 159/181 (87.8%) | 87/96 (90.6%) | 72/85 (84.7%) |  |
| Syphilis test result |  |  |  | >0.999 |
| Negative | 272/273 (99.6%) | 125/125 (100.0%) | 147/148 (99.3%) |  |
| Positive | 1/273 (0.37%) | 0/125 (0%) | 1/148 (0.68%) |  |
| HIV test result |  |  |  | 0.448 |
| Negative | 323/329 (98.2%) | 160/164 (97.6%) | 163/165 (98.8%) |  |
| Positive | 6/329 (1.82%) | 4/164 (2.44%) | 2/165 (1.21%) |  |

**Table S4. Description of suspected mpox cases by PCR result**

| **Characteristic** | **Overall** (n = 415) | **Negative** (n = 93) | **Positive** (n = 322) | **p-value** |
| --- | --- | --- | --- | --- |
| Sex |  |  |  | 0.992 |
| Female | 201/415 (48.4%) | 45/93 (48.4%) | 156/322 (48.4%) |  |
| Male | 214/415 (51.6%) | 48/93 (51.6%) | 166/322 (51.6%) |  |
| Age in years, median (IQR) | 9 (2.8, 21.0) | 7 (2.0, 16.0) | 10 (3.0, 22.0) | 0.134 |
| Age group |  |  |  | 0.213 |
| Under 1 year | 65/415 (15.7%) | 15/93 (16.1%) | 50/322 (15.5%) |  |
| 1-4 years | 90/415 (21.7%) | 22/93 (23.7%) | 68/322 (21.1%) |  |
| 5-14 years | 114/415 (27.5%) | 32/93 (34.4%) | 82/322 (25.5%) |  |
| 15-44 years | 141/415 (34.0%) | 23/93 (24.7%) | 118/322 (36.6%) |  |
| ≥ 45 years | 5/415 (1.20%) | 1/93 (1.08%) | 4/322 (1.24%) |  |
| Hospitalized | 238/395 (60.3%) | 39/85 (45.9%) | 199/310 (64.2%) | 0.002 |
| Self-reported fever | 322/388 (83.0%) | 67/83 (80.7%) | 255/305 (83.6%) | 0.535 |
| Lesions on palms and soles | 172/397 (43.3%) | 27/87 (31.0%) | 145/310 (46.8%) | 0.009 |
| Cervical adenopathy | 289/399 (72.4%) | 57/86 (66.3%) | 232/313 (74.1%) | 0.149 |
| Inguinal adenopathy | 340/399 (85.2%) | 75/86 (87.2%) | 265/313 (84.7%) | 0.556 |
| Axillary adenopathy | 171/398 (43.0%) | 27/86 (31.4%) | 144/312 (46.2%) | 0.014 |
| Generalized adenopathy | 131/382 (34.3%) | 20/82 (24.4%) | 111/300 (37.0%) | 0.033 |
| Eye redness | 62/394 (15.7%) | 10/85 (11.8%) | 52/309 (16.8%) | 0.256 |
| Eye itching | 45/227 (19.8%) | 7/47 (14.9%) | 38/180 (21.1%) | 0.341 |
| Light sensitivity | 50/218 (22.9%) | 7/45 (15.6%) | 43/173 (24.9%) | 0.186 |
| Oral lesions | 125/392 (31.9%) | 16/87 (18.4%) | 109/305 (35.7%) | 0.002 |
| Genital lesions | 221/396 (55.8%) | 45/86 (52.3%) | 176/310 (56.8%) | 0.462 |
| Anal lesions | 62/392 (15.8%) | 8/86 (9.30%) | 54/306 (17.6%) | 0.061 |
| Number of lesions |  |  |  | 0.400 |
| <25 lesions | 99/395 (25.1%) | 25/85 (29.4%) | 74/310 (23.9%) |  |
| 25-99 lesions | 200/395 (50.6%) | 45/85 (52.9%) | 155/310 (50.0%) |  |
| 100-250 lesions | 68/395 (17.2%) | 11/85 (12.9%) | 57/310 (18.4%) |  |
| >250 lesions | 28/395 (7.09%) | 4/85 (4.71%) | 24/310 (7.74%) |  |
| Pregnancy status | 8/61 (13.1%) | 2/10 (20.0%) | 6/51 (11.8%) | 0.607 |
| Nutritional status of children |  |  |  | 0.488 |
| Severe acute malnutrition | 4/96 (4.17%) | 0/24 (0%) | 4/72 (5.56%) |  |
| Moderate acute malnutrition | 5/96 (5.21%) | 2/24 (8.33%) | 3/72 (4.17%) |  |
| Normal | 87/96 (90.6%) | 22/24 (91.7%) | 65/72 (90.3%) |  |
| Syphilis test result |  |  |  |  |
| negative | 125/125 (100.0%) | 28/28 (100.0%) | 97/97 (100.0%) |  |
| HIV test result |  |  |  | 0.577 |
| Negative | 160/164 (97.6%) | 46/46 (100.0%) | 114/118 (96.6%) |  |
| Positive | 4/164 (2.44%) | 0/46 (0%) | 4/118 (3.39%) |  |

### 3. **Pregnancy status in the Mpox Treatment Center in Uvira**

Between June 3 and October 24, 2024, we recorded 131 women of childbearing age (15-49 years), of whom 19 reported to be pregnant (Table S5). No pregnancy test was offered in the MTC at the time of this study.

Of the 19 pregnant women included in the study, follow-up home visits conducted by Uvira Hospital were successfully completed for 10 participants, four of whom had tested positive for mpox during hospitalization at the MTC. Among the remaining nine, two resided outside Uvira city, and seven had unknown addresses, preventing further contact. At the time of manuscript preparation (mid-December 2024), eight of the ten followed-up women had delivered, resulting in nine live births, including one set of twins. Birth weights ranged from 2,500 g to 4,000 g. The remaining two women were still pregnant. All deliveries were uneventful, with no reported neonatal complications. All newborns were in good health, and no rash lesions were observed by the mothers. Additionally, none of the ten pregnant women reported obstetric complications during pregnancy.

**Table S5. Description pregnant women in the Uvira Mpox Treatment Center**

| Characteristic | N = 19 |
| --- | --- |
| Age | 21 (19.0, 28.0) |
| Gestational age |  |
| Q1 | 3/19 (15.8%) |
| Q2 | 5/19 (26.3%) |
| Q3 | 7/19 (36.8%) |
| Unknown gestational age | 4/19 (21.1%) |
| Hospitalized | 13/19 (68.4%) |
| Self-reported fever | 13/19 (68.4%) |
| Presence of genital lesions | 12/19 (63.2%) |
| Presence of anal lesions | 3/19 (15.8%) |
| Number of lesions |  |
| <25 lesions | 7/19 (36.8%) |
| 25-99 lesions | 9/19 (47.4%) |
| 100-250 lesions | 1/19 (5.26%) |
| >250 lesions | 2/19 (10.5%) |
| Absence of vaginal bleeding | 17/17 (100.0%) |
| Absence of uterine contractions | 16/16 (100.0%) |
| Negative syphilis test result | 6/6 (100.0%) |
| HIV test results |  |
| Negative | 6/7 (85.7%) |
| Positive | 1/7 (14.3%) |
| Mpox confirmation (PCR) |  |
| Positive | 6/8 (75.0%) |
| Negative | 1/8 (12.5%) |
| Indeterminate | 1/8 (12.5%) |

Variations in the denominator reflect differences in data availability for each characteristics.

### 4. **Description of mpox deaths at the Uvira Mpox Treatment Center**

At the Uvira MTC, clinical outcomes for all suspected mpox cases were documented on discharge forms. In cases of death, information on clinical progression, length of hospitalization, treatments administered, complications, and probable causes of death was extracted from medical records. During the study period, seven deaths were reported: five in infants under one year of age and two in immunocompromised adults. All fatal cases presented with respiratory distress and bacterial superinfection, requiring oxygen therapy prior to death. Additionally, three cases developed anemia during hospitalization and required blood transfusions. Detailed clinical descriptions of the fatal cases are provided in Table S6.

**Table S5. Brief description of suspected mpox deaths in Uvira**

| **Case** | **Description** |
| --- | --- |
| **Case 1** | A 30-35-year-old female sex worker was admitted in mid-June 2024 to the Uvira Mpox Treatment Center (MTC) with severe and disseminated eruptive skin lesions. She reported the onset of skin lesions and fever four days prior to admission. On arrival, she exhibited disseminated vesiculopapular lesions, erythema, and pustular exacerbations on her palms, anal area, and genital region, along with inguinal lymphadenopathy. Her HIV status was unknown prior to admission but was confirmed as HIV-positive via rapid diagnostic tests (RDT), while a syphilis RDT was negative. Mpox PCR test result was not available. She also presented with hyperglycemia and was diagnosed with diabetes during her hospital stay. She was admitted to the intensive care unit (ICU) for severe coma (Glasgow coma scale 3/15) and received probabilistic empirical antibiotic therapy (ceftriaxone, ciprofloxacin, and metronidazole), intravenous fluids (NaCl 0.9%), insulin, oxygen therapy, and wound care. Throughout her ICU stay, she developed complications, including acute respiratory distress and renal failure. She died three days after admission due to uncontrolled diabetes complicated by hyperosmolar coma and suspected severe mpox with a respiratory bacterial superinfection. |
| **Case 2** | A female patient, less than one month old was admitted to the Uvira MTC in late July 2024 with suspected severe mpox. Two days prior, she developed a disseminated rash and fever. Upon admission, she presented with generalized superficial lymphadenopathy and disseminated macular skin lesions, including on her genitals and palms. Her weight was 5 kg. Length measurement was unavailable due to a lack of appropriate tools. The patient was born full-term following an uncomplicated pregnancy and delivery, with no abnormalities or skin lesions observed at birth. Neither her mother nor other household members exhibited signs of mpox. The mother self-reported being healthy, HIV-negative, and having an uneventful delivery. Skin lesion samples collected at admission tested positive for mpox using PCR. During hospitalization, the patient developed severe complications, including a skin bacterial superinfection, acute respiratory distress, and anaemia, in a context of sepsis. She received antibiotics (ceftriaxone), oxygen therapy, a blood transfusion in the intensive care unit, and treatment with sulfadiazine ointment, paracetamol, and vitamin A. Despite intensive care, she succumbed to acute respiratory distress and anaemia on day 24 of hospitalization. |
| **Case 3** | An 10-12-month-old female patient was hospitalized at the Uvira MTC in early August 2024 with suspected severe mpox. Symptoms began six days prior to admission, starting with high fever followed by a rash four days later. Upon admission, she presented with papular, macular, and erythematous lesions on her soles, palms, and perineal area. No other family members exhibited signs of mpox. She had inguinal lymphadenopathy, weighed 7 kg, and had an arm circumference of 134 mm. Mpox qPCR testing of her skin lesions was positive, while HIV and syphilis RDTs were negative. During hospitalization, she developed skin bacterial superinfections, respiratory distress, and sepsis. She was treated with ceftriaxone, sulfadiazine ointment, paracetamol, and vitamin A, and received oxygen therapy in the intensive care unit. She passed away due to acute respiratory distress on day 21 of hospitalization. |
| **Case 4** | An 10-12-month-old male patient was hospitalized at the Uvira MTC in early August 2024 with suspected severe mpox. Symptoms began three days earlier with fever, followed by the development of a rash two days later. Upon admission, he presented with disseminated papular skin lesions covering his entire body, including the oropharyngeal region (tongue, gums, soft palate) and genitals, along with generalized superficial lymphadenopathy. He weighed 11 kg, and his arm circumference measured 141 mm. Three days after his admission, his mother exhibited signs of mpox, and her HIV serological status was unknown. Testing of the patient's skin lesions using qPCR was negative, and both HIV and syphilis RDTs were also negative. During his hospitalization, he developed a skin bacterial superinfection, respiratory distress, and sepsis. He was treated with ceftriaxone, sulfadiazine ointment, paracetamol, and vitamin A, and received oxygen therapy and a blood transfusion. One week after admission, the patient passed away due to acute respiratory distress associated with anemia in the context of sepsis. |
| **Case 5** | A 30-35-year-old male nurse, HIV-positive, was referred from a remote rural hospital to the Uvira MTC. Upon admission, he reported respiratory discomfort, high-grade fever, disseminated ulcero-purulent skin lesions, generalized superficial lymphadenopathy, bilateral purulent otorrhea, and significant psychological distress, including suicidal thoughts. Mpox was confirmed by qPCR testing. A detailed case report of this patient is presented elsewhere.[[1]](https://paperpile.com/c/NGEtQA/9rDL) |
| **Case 6** | A 0-3-month-old female patient was hospitalized at the Uvira MTC in early October 2024 with suspected severe mpox. Symptoms began seven days prior to admission with a disseminated skin rash, followed by fever four days later. Upon admission, she presented with multiple papular lesions scattered across her body, including her palms, soles, and genitals, as well as oral lesions primarily affecting her tongue. Her weight was 5 kg, and her arm circumference measured 140 mm. The patient exhibited coughing, rhinorrhea, and persistent fevers, with temperatures reaching 38.5°C. Generalized superficial lymphadenopathy was also noted. Her mother did not show signs of mpox, and her HIV serological status was unknown. The treatment regimen included ceftriaxone, cloxacillin, paracetamol, and vitamin A, along with sulfadiazine ointment for lesion dressings. The patient received oxygen therapy and nystatin in the intensive care unit. On Day 5 of hospitalization, she passed away due to acute respiratory distress. The mpox qPCR test result was positive. |
| **Case 7** | A 0-3-month-old male patient was admitted to the Uvira MTC in early October 2024 with suspected severe mpox. Symptoms began four days prior with fever and lymphadenopathy. Upon admission, he exhibited vesicular, umbilicated pustular, and macular lesions covering his entire body, including his palms, soles, oropharyngeal region (tongue, gums, and palate), and genitals. He came from a family of seven, none of whom had shown signs of mpox, and his mother's HIV serological status was unknown. The patient also presented with cough, diarrhea, and respiratory distress. His treatment included ceftriaxone, cloxacillin, paracetamol, loratadine, and vitamin A. He passed away three days after admission due to acute respiratory distress and dehydration. The mpox qPCR testing of his skin lesions was positive. |

MTC: Mpox Treatment Center.

### **References**

1.	Bugeme PM, Bugale PK, Mashauri JZ, et al. Disseminated mpox in an immunocompromised patient in DR Congo: a call for a shift from disease-focused to person-centred outbreak response. Lancet Infect Dis **2025**; Available at: <http://dx.doi.org/10.1016/s1473-3099(25)00009-x>.
